# criTRia: A Classification System and Evidence Criteria for Tandem Repeat Locus-Disease Relationships

**DOI:** 10.64898/2026.07.04.26357279

**Authors:** Macayla Ann Weiner, Laurel Hiatt, Pamela Ajuyah, Elbay Aliyev, Harriet Dashnow

## Abstract

**Introduction:** Tandem repeats (TRs), including short tandem repeats (1-6 bp motifs) and variable number tandem repeats (7+ bp motifs), have been linked to more than 50 Mendelian diseases. However, current frameworks for evaluating gene-disease relationships do not adequately address TR-specific complexities. As a result, proposed TR locus-disease relationships are often incorrectly classified, under-evaluated, or excluded entirely, limiting discovery and leading to underdiagnosis of TR disorders.

**Methods:** We developed criTRia, a scoring framework designed to accurately evaluate TR locus-disease relationships at the locus level rather than the gene level. Building on ClinGen best practices, criTRia introduces TR-specific evidence categories and reweighted scoring. We applied criTRia to curate 65 loci from STRchive, a database of disease-associated TRs.

**Results:** We compared criTRia curations with gene-level curations from nine Gene Curation Coalition (GenCC) groups. Of 65 newly scored loci, 7 had not been previously evaluated by GenCC and 17 showed significant disagreement across groups. These differences have direct implications for whether a disease is recommended for inclusion in a diagnostic gene panel. The criTRia framework also enabled curation of previously unassessed associations, bringing the total to 77 curated TR locus-disease associations and identifying four contradictory associations.

**Discussion:** By incorporating TR-specific evidence, criTRia provides a reproducible methodology for assessing TR locus-disease relationships, improving classification consistency and establishing a foundation for better integrating tandem repeats into clinical genetic medicine and providing more accurate diagnoses.

## Introduction

### The problem and its importance

Accurate curation of gene-disease relationships is foundational for reliable clinical interpretation and advancing rare disease diagnostics. These curations guide diagnostic variant interpretation, support genetic counseling, and provide a framework for rare disease research^1^. Consortia such as ClinGen^2^ have established standardized approaches to evaluating evidence^3^, supporting consistency and transparency across laboratories and institutions. These entities, along with other global initiatives like Genomics England PanelApp and Orphanet, as well as diagnostic laboratories such as Ambry Genetics and Labcorp/Invitae, are member organizations of the Gene Curation Coalition (GenCC)^4^ which harmonizes their terminology so that curations from different institutions can be compared in a unified database. Yet these efforts have predominantly focused on interpreting small coding variants, including single-nucleotide variants (SNVs), short insertions/deletions (indels), and copy number variations (CNVs), as well as large structural variants, such as chromosomal-level rearrangements^5–7^. 1 in 3000 people are predicted to have tandem repeat (TR) diseases, with the majority undiagnosed^8^. TR diseases have often been overlooked in clinical diagnostics due to their complexity.

TRs, including short tandem repeats (STRs, 1-6 bp motifs) and variable number tandem repeats (VNTRs, 7+ bp motifs), have been implicated in more than 50 Mendelian diseases such as the spinocerebellar ataxias and Huntington’s disease^9^. Despite frequent involvement of TR loci in disease, there has been little guidance or standardization for determining whether newly reported TR disease loci are, in fact, clinically relevant. We previously created STRchive^10^, a central repository for all Mendelian TR locus-disease relationships. During the development of STRchive, it became clear that a new scoring procedure was necessary to indicate the level of evidence for a given locus-disease association. While numerous scoring guidelines^2,5,6,11–17^ exist, in practice, none have adapted well to TR diseases, as evidenced by the groups’ scoring histories, leaving us with incomplete capture of reported TR diseases. For example, there are 23 gene-disease relationships for TR loci curated by ClinGen’s working groups reported on the Gene Curation Coalition^4^ database, 19 of which have a “Definitive” or “Strong” score, and very few of which have been re-evaluated, indicating that only well-established TR disease loci are chosen for curation. With the rapid expansion of TR research and continued advances in sequencing and computational methods^18–21^, the rate of TR discovery has far outpaced curation and clinical validation efforts^22^. Many existing curation procedures tend to wait for substantial evidence before publishing their classifications. Part of the reason is that TRs possess unique attributes that challenge current curation strategies.

### What is special about TR diseases?

A clear need has emerged for tandem-repeat-aware scoring criteria, as these conditions have nuances absent in other genetic diseases. For example, current gene-level scoring approaches do not allow us to directly consider tandem repeat-specific information such as motif composition or repeat length consequences. Furthermore, our ability to computationally predict pathogenicity for TR loci is limited and differs from metrics used for other variant types, while population allele frequency data has only recently become available^19,23^. Examples of criteria we have adapted from ClinGen to better score TR diseases include the scoring for variant evidence and the definition of segregation evidence. Additionally, we endeavor to address putative TR loci as evidence becomes available, acting at the pace that is necessary to include TR diseases in contemporary clinical diagnostics. Our approach builds on proven ClinGen curation practices^3,24^ while extending them to address the nuances of TR diseases.

In cases of repeat-specific diseases, scoring at the gene level is typically insufficient and may oversimplify evidence classifications. Familial adult myoclonic epilepsy (FAME) illustrates this complexity well. Rather than using a gene to identify the disease relationship, researchers observed the same motif (TTTCA) in many different genes^25^. Researchers were able to causally identify this motif-disease relationship without a prior gene-disease relationship. Had researchers only been looking at the gene-level, they never would have identified that FAME subtypes on different genes were related. Moreover, some genes have both pathogenic and apparently benign repeat tracks or multiple distinct pathogenic tracks, such as *HOXA13*^*26*^. Duchenne muscular dystrophy (DMD) is another example in which locus-rather than gene-level specificity is crucial for characterizing TR disease. One 2016 study proposed that a TR locus within the *DMD* gene may lead to a DMD phenotype^27^; although the *DMD* gene is known to underlie the DMD disease and there is substantive evidence at the gene-level, later population-level evidence demonstrated that the distribution of repeat sizes at this locus reflects ancestry and is unrelated to disease phenotype^18,28^. Creating a resource to score evidence based on the TR locus ensures the sensitivity and specificity required in evaluating this variant type at scale.

TR disease mechanisms are distinct. Unlike other variant types, the allele size and motif structure of TRs can allow for discrete, repeat-based disease mechanisms. Previous gene-level scoring frameworks, which rely on assumptions derived from other variant classes, do not fully capture the range of these TR-specific mechanisms. Repeats can form abnormal RNA structures that sequester binding proteins or be translated via repeat-associated non-AUG (RAN) translation into neurotoxic peptide chains. These effects often scale with repeat length and involve both coding and noncoding regions, leading to toxic protein products. New research has introduced TR-specific evaluative measures, such as constraint and longest pure motif length, to prioritize potential pathogenic sites. Repeat-specific software, such as STRipy^20^, has been introduced to accurately identify unique features of TRs. With repeat-aware methods already in practice for evaluating candidate loci, there is a need for TR-specific scoring that accounts for these factors.

Additionally, TR diseases exhibit clinical features that distinguish them from other variant types. Unlike other variants, repeat motif and length can dramatically influence disease phenotype, including age of onset and severity. For example, longer alleles often correlate with earlier onset and more severe clinical presentation, while shorter alleles or interrupted motifs may modulate expressivity or reduce penetrance (such as Huntington’s disease on *HTT*^*29*^ and Friedreich’s Ataxia on *FXN*^*30*^). Variations in allele size, interruptions, mosaicism, and methylation can all influence age of onset and penetrance, complicating clinical interpretation^22^.

Given that TRs are still relatively emergent in genetics research, there is much to learn as research continues to evolve. Current classifications calibrated for non-TR entities are updated too infrequently to keep pace with emerging data in the TR space. While iterating on our scoring criteria, we observed scores change from “Limited” to “Definitive” and reach the maximum numerical score solely because substantial research was published over a short period. For this reason, we adjusted our re-scoring time frames from ClinGen norms; the “Limited” category will be rescored every year instead of every three, and “Strong” every two years instead of every three.

### The Solution

Here, we present criTRia, a protocol for documenting and scoring TR loci associated with Mendelian disease that evaluates the strength of locus-disease relationships using publicly available evidence. The criTRia framework adapts and extends ClinGen’s gene-disease relationship standard operating procedure^31^ to specifically address and incorporate TR-specific criteria and considerations. Future researchers may also use criTRia to improve clarity and ensure accuracy of reported data, facilitating integration into diagnostic pipelines and future research efforts. This will improve collaboration across labs and institutes while developing more robust frameworks for these diseases. Information about the locus-disease relationship, including genetic, experimental, and contradictory evidence scored from publicly available sources, is compiled and used to assign a numeric (0-18) and categorical (“Definitive”, “Strong”, “Moderate”, “Limited”, “No Known” TR locus-disease relationship, “Disputed”, and “Refuted”) score to the locus-disease relationship per established criteria. Our protocol outlines the steps for scoring a locus-disease relationship and assigning a validity classification. This process involves an overview of the pertinent evidence available to assign the appropriate classification for a locus-disease relationship at a given time. While the following protocol provides guidance on the scoring process and our classifications are reported in the resource STRchive, professional judgment and expertise, where applicable, must be used when interpreting evidence that supports a locus-disease relationship.

Accurate scoring of even the least contextualized loci is essential, especially in clinical settings. An unfounded relationship with no score or the wrong score can lead to missed diagnoses, incorrect diagnoses, delayed treatment, and unnecessary genetic counseling. Beyond clinical settings, systematically evaluating all known loci will help researchers identify novel pathogenic mechanisms, refine our understanding of these relationships, and evolve as methodologies develop. Standardized classification facilitates population-level analysis, which in turn can pave the way for the identification of complex traits.

## Methods

We identified 76 TR loci for assessment from STRchive v2.22.0, excluding four provisional loci with insufficient evidence for scoring. For each locus, we first checked whether ClinGen had performed gene-level curation and then assessed it against the criTRia protocol, identifying 12 loci that had been well-scored by ClinGen. We then proceeded to fully curate the remaining 65 loci. This 65 included two entries for *FMR1*, which was split into diseases FXS and FXTAS/POF1 by repeat length. The full curation and scoring procedure is described in the criTRia SOP, version 1 (**Supplemental File 1**). Here, we briefly summarize the process for the curations performed as part of this publication.

### Numerical Scoring

The evidence scored came from peer-reviewed publications and high-quality preprints, primarily using literature cited in STRchive. Evidence was categorized as “Genetic” and “Experimental” (Figure 1), with maximum locus scores of 12 and 6, respectively, and pieces of evidence assigned to specific sections within each category. When looking for genetic evidence, proband/inheritance evidence, repeat length or motif consequences, method of predicting pathogenicity, segregation evidence, and statistical significance were collected and evaluated. Scores ranged from 0-18 (0 being no evidence and 18 being the maximum amount of evidence for a “Definitive” classification if evidence was repeated over time). 65 locus-phenotype relationships were scored and classified based on current available evidence.

**Figure 1.**
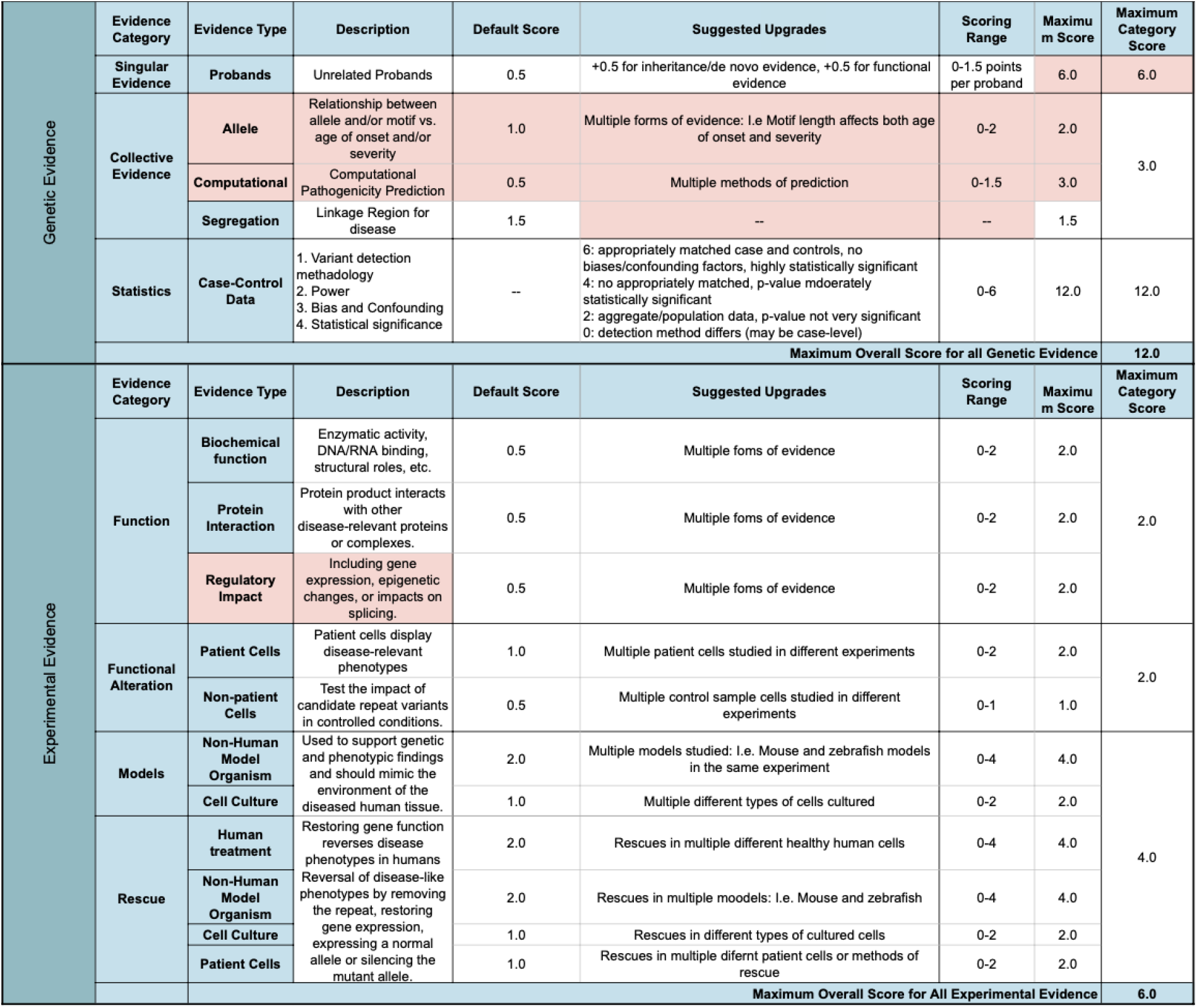
Evidence Matrix, highlighting in red differences from the ClinGen Gene Curation framework for TR-specificity.

### Categorical Scoring

From the numerical scores, we binned the loci into categorical scores, “Definitive”, “Strong”, “Moderate”, “Limited”, “No Known” TR locus-disease relationship, “Disputed”, and “Refuted”. Scores between 12 and 18, where evidence has been repeated over 3 years, receive “Definitive”. Otherwise, these loci are classified as “Strong”, while loci with 7-11 points receive “Moderate”, and those with fewer than 7 receive “Limited”. The “No Known” TR locus-disease relationship category is given when there has been no evidence relating the TR to a disease. Finally, there are two conflicting categories. “Disputed” is designated when the relationship has been disproven, cases have disparate phenotypes, or where disease appears polygenic rather than Mendelian, after being reported as Mendelian. This category comes with the caveat that no alternative cause for the disease has been identified. If there has been an alternative cause, the relationship will likely fall under the “Refuted” category. However, this designation may also arise from other lines of evidence, such as the re-evaluation of previously reported variants as too frequent in the general population to be disease-causing. Loci may also be assigned this category if all evidence has been ruled out or if the proband has been proven to have a different disease.

### Adding evidence detail

Disease-locus relationships were first curated by manually reviewing the relevant literature and scoring each evidence category as described in the criTRia SOP, version 1 (**Supplemental File 1**). A large language model was used to generate a concise summary of the evidence in the paper for each score and a locus-level summary. This text was generated using a semi-automated workflow with ChatGPT (version GPT-5.5 Thinking) assistance^32^. A standardized prompt was used across entries to ensure consistency in the generation of evidence details (see **Supplemental File 2)**. ChatGPT was used only to draft concise evidence-detail descriptions from the provided source materials and structured criTRia entries; evidence scores, categories, summary scores, classifications, and schema structure were not generated or modified by ChatGPT. All AI-assisted evidence-detail text was manually reviewed against the source literature and approved by at least one additional human reviewer before inclusion. Additionally, the prompt included a request to assess each score and paper against the SOP and note any inconsistencies. Each statement was then reviewed by a different curator for both accuracy and relevance of that evidence to the overall curation. At least two curators, typically three, curated each locus.

## Results

We identified 76 TR loci for assessment from STRchive v2.22.0, excluding four recently added provisional loci with insufficient evidence for scoring. One locus (FMR1) was split to have two curations (FXS and FXTAS/POF1). Twelve gene-disease relationships had previously been scored “Definitive” by ClinGen (ADTKD_MUC1, CANVAS_RFC1, CCHS_PHOX2B, EDM1-PSACH_COMP, EPM1_CSTB, FRAXE_AFF2, FRDA_FXN, FTDALS1_C9orf72, GDPAG_GLS, HD_HTT, SBMA_AR, SCA2_ATXN2). We reviewed these gene-level curations and determined that they did not need to be re-evaluated at the locus level, as they were already strongly supported and incorporation of additional TR-specific evidence was unlikely to affect the classification outcome. Our final dataset (**Supplemental File 3**) contained 77 TR locus-disease relationships and 76 unique loci, 65 of these curated using the criTRia framework (detailed curations in **Supplemental File 4**).

### Strength of Evidence Varies Across Loci

After curation using the criTRia framework, the majority of loci were classified as “Definitive” or “Strong” (39/65, 60%), with a further 8 classified as “Moderate” (12%). Smaller proportions were categorized as “Limited” (14/65, 22%) and “Contradictory” (“Disputed” or “Refuted”, 4/65, 6%) (Figure 2). This distribution demonstrates that although many TR disorders are strongly supported, a substantial subset remains incompletely or incorrectly characterized.

**Figure 2.**
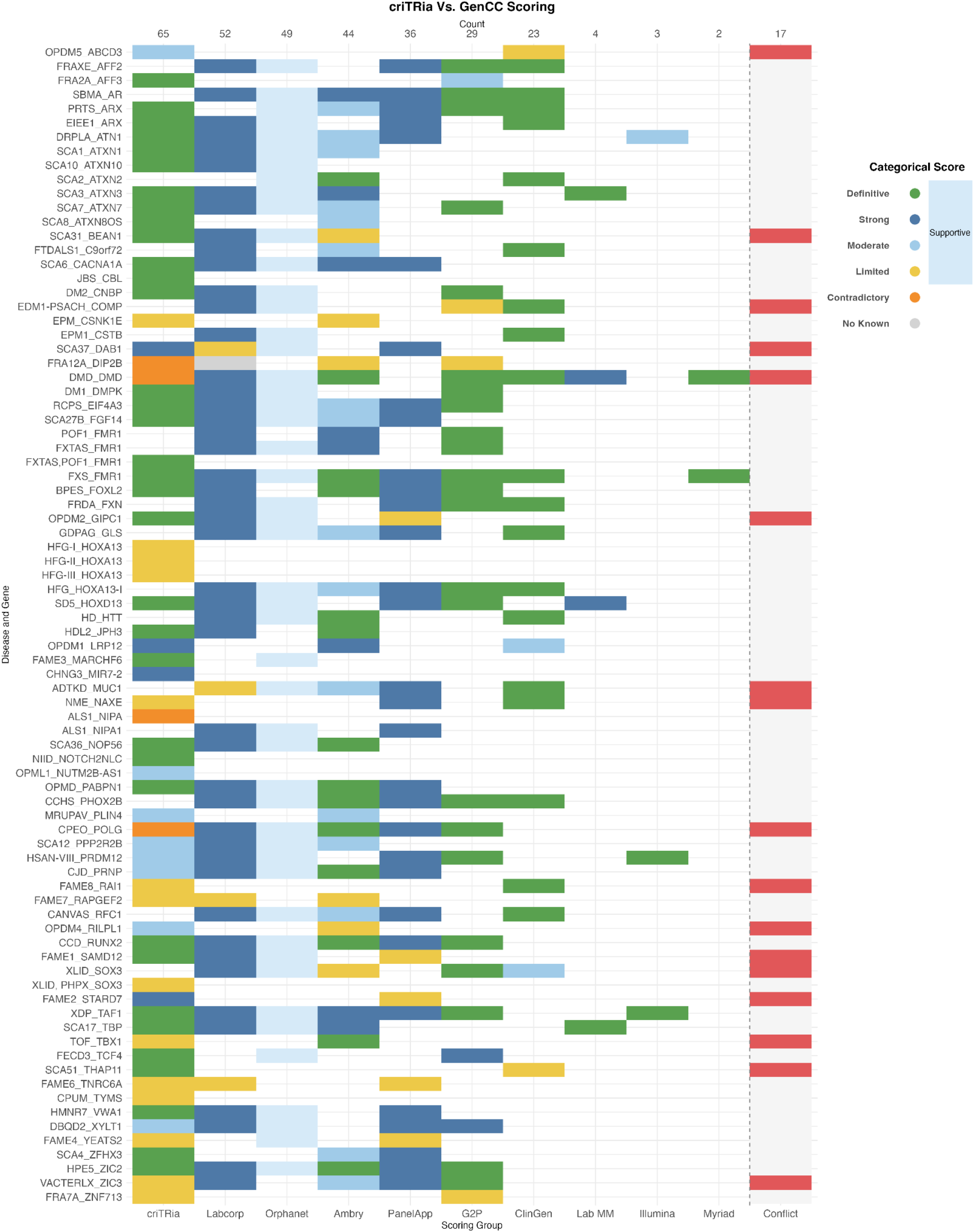
Comparison of scoring across curating groups: Rows represent disease-gene or disease-locus relationships and columns represent GenCC-submitting curation groups, including Labcorp, Orphanet, Ambry, PanelApp, G2P, ClinGen, Lab MM, Illumina, and Myriad. Colors indicate categorical classification strengths ranging from “Contradictory” to “Definitive”. The “Supportive” used by Orphanet overlaps with all positive categories: Limited, Moderate, Strong, and Definitive. The criTRia classifications spanned the full range of evidence categories, while the majority of scores from GenCC submitters tended to fall into higher-confidence categories such as “Strong” or “Definitive”. Well-established repeat expansion disorders typically showed agreement. Differences in category usage reflect variability in curation frameworks between submitting groups. The “Conflict” column indicates loci in red where at least one group assigned a high-evidence classification (Definitive, Strong, or Moderate) and at least one other assigned a low-evidence classification (Limited, Contradictory, or No Known). The Supportive category was not considered for categorizing Conflict.

### Comparisons to Gene-level Frameworks

We compared criTRia classifications with those from several Gene Curation Coalition (GenCC) members, including ClinGen, Ambry Genetics, and Orphanet (Figure 2). Of the 80 we report on STRchive, 59 were in genes already reported in the GenCC database^4^. ClinGen had produced gene-level curations for 23 of these, and the majority were scored as “Definitive” or “Strong” (19/23, 73%). None were reported as “Conflicting”. This comparison shows that gene-level and locus-level approaches are often consistent for well-established disorders, but differences still emerge when repeat-specific mechanisms are considered.

In total, 52 relationships showed consistent classifications across frameworks, whereas inconsistent cases, including Duchenne Muscular Dystrophy (DMD), progressive ophthalmoplegia (CPEO), and hand-foot-genital syndromes (HFG1-3), demonstrated how gene-level scoring may obscure distinct repeat loci within the same gene. For example, HFG1-3 were evaluated as separate locus-disease relationships because the pathogenic coordinates differed within the gene, distinctions not captured in gene-level frameworks.

Of the genes scored by multiple groups, 17 showed discordant evidence tier classifications: at least one group assigned a positive evidence category (Definitive, Strong, or Moderate), while at least one other assigned a low-evidence category (Limited, Contradictory, or No Known).

This distinction is clinically meaningful: positive classifications support inclusion in diagnostic gene panels, whereas low-evidence classifications indicate associations that remain in the research domain^33,34^. These discordances highlight genes where the clinical interpretation of a TR locus-disease relationship differs meaningfully depending on the curation framework applied.

### Evaluation of Previously Uncurated Associations

Of the 7 previously uncurated TR locus-disease associations, many are not well supported (“Limited”: 3) or have different evidence profiles from those used to support the broader gene-disease relationship: e.g., *DMD* as a gene has strong clinical associations but does not have disease evidence associated with the TR locus. Experimental evidence often appeared more abundant than genetic evidence across these loci, despite still presenting ambiguous clinical support, emphasizing the need for balanced weighting between mechanistic and human genetic data. These evaluations additionally identified four conflicting relationships and clarified several weakly supported or disputed associations, enabling more consistent interpretation of emerging TR disease candidates.

### Curating Challenging Loci

We gave special care to cases where multiple TR loci are reported in the same gene. Many of these loci have only recently been defined, and, in turn, much of the current literature may not be as robust as it appears at the gene level. Occasionally, evidence of a broad phenotypic spectrum must be evaluated, where distinctions between loci may be inferred from features such as symptom presentation or age of onset.

### Subtypes of the Same TR-Disease

Some genes contain multiple TR-diseases or subtypes of a disease on the same gene; these subtypes generally represent a spectrum of a single disease rather than separate entities.

However, in some cases different diseases may be defined by repeat size, composition, inheritance pattern, or severity of presentation. The following is an example of how we scored such diseases:

#### Hand-foot-genital (HFG) syndrome of HOXA13

This phenotype has been linked to TR expansions in three distinct polyalanine tracks, each separated by ∼60 base pairs. This has led to the proposed three conditions: HFG-I, HFG-II, and HFG-III associated with the loci *HOXA13-I, HOXA13-II*, and *HOXA13-III*, respectively. The proximity and similarity of these loci make it difficult to accurately and confidently score the evidence. The pathogenic range of the repeats provided unique “landmarks” for each disease subtype. For most papers, this gave us a clear indication of the type of HFG the publications were referring to. Any papers that were still unclear were scored across all three types, and restricted to the experimental category as these were typically gene-level experiments.

### Multiple TR-Diseases Associated with the Same Gene

In other cases, distinct TR-disease may arise from different repeat ranges, motifs, or loci within the same gene. These should generally be considered as separate diseases when they produce distinguishable phenotypes or mechanisms. The following are two examples of how we scored cases like this:

#### Diseases of FMR1

Fragile X syndrome (FXS), fragile X-associated tremor/ataxia syndrome (FXTAS), and fragile X-associated premature ovarian failure (POF1) all exist as a spectrum of disease within the same repeat locus in *FMR1*. This was a unique problem, as FXTAS and POF1 are associated with the intermediate expansion range (55-200 repeats), and FXS occurs at the “full” mutation (>200 repeats), and some symptoms may overlap. Identifying FXS-focused papers was fairly straightforward, as most explicitly defined the repeat range. It became increasingly clear that FXTAS and POF1 were entangled within the literature, as there were overlapping symptoms in XX (genetically female) individuals with both diseases, and the bioinformatic evidence was similar, and maybe identical. For this reason, FXTAS and POF1 were combined, and FXS was scored separately^35^.

#### The ARX phenotypic spectrum

Expansions in *ARX* vary in length and location on the gene and cause a range of diseases. For this curation, we focused on Partington syndrome (PRTS) and early infantile epileptic encephalopathy (EIEE1), as these are the two conditions that have been associated with TRs. Many studies examine all ARX diseases in aggregate and do not clearly define which disease is being represented when presenting the evidence. Most studies were easily linked to one disease, but for those that were not, it was decided that this evidence would be scored in the experimental category for both conditions if findings were relevant to both phenotypes. It is worth noting that while locus coordinates are usually a useful tool for distinguishing TR diseases, it was not advantageous in this specific situation.

## Discussion

Despite the growing recognition of the importance of TRs in human disease^22^, scoring frameworks capable of precisely evaluating TR loci have been lacking. Most existing frameworks take a gene-level view and do not account for the nuanced, context-specific behavior of TRs. As a result, some evidence has been under- or mis-evaluated, leaving potentially pathogenic loci overlooked or misclassified. This limitation is reflected in our cross-framework comparison: while criTRia classifications broadly correspond with scores from other groups for well-established loci, the vast majority of TR disease genes scored by non-TR-focused groups carry a “Definitive” classification. This pattern reveals a systemic bias: these frameworks selectively curate only genes containing the best-characterized TR disease loci, leaving more uncertain or under-researched repeats unaddressed and beyond the reach of existing diagnostic frameworks.

By systematically evaluating even poorly characterized loci and integrating genetic and experimental evidence in a locus-specific scoring framework, our approach fills a critical gap in the field. This gap is especially consequential in clinical settings, where accurate interpretation of locus clinical relevance is crucial in variant prioritization and clinical diagnosis. The 17 loci showing tier-level discordance illustrate this directly: for these loci, the choice of curation framework may determine whether the associated disease is included in a diagnostic gene panel or remains confined to research settings^33,34^. A consistent, locus-aware approach is therefore not just an academic distinction but a clinical one. The lack of scoring procedures specific to locus-level diseases also affects locus discovery and population-level analyses.

Large-scale catalogs and functional studies generate unprecedented amounts of research, yet we have no dedicated scoring procedures to parse these new data. Without TR-focused strategies, we cannot leverage this research to accurately identify novel repeats or refine genotype-phenotype relationships. By creating a repeat-specific, evidence-based scoring system, our work enables both clinical and research communities to extract meaningful insights from TR data that might otherwise go unassessed.

### Minimum Evidence when publishing a new TR locus-disease association

We strongly urge researchers publishing a novel locus-disease relationship to provide sufficient detail and evidence to allow future genetic testing and curation. We outline our recommended genetic and bioinformatic evidence in **Figure 3**. First, any publication describing a TR variant should specify the reference genomic coordinates with as much accuracy as possible, given the discovery method used. For studies using high-resolution methods such as DNA sequencing or PCR, single bp or near-bp accuracy is expected. Statements like “expansion in the first intron of gene X” are insufficient to develop an accurate diagnostic assay and can lead downstream users to reconstruct genomic coordinates from paper figures and supplementary data, potentially resulting in inconsistencies.

**Figure 3.**
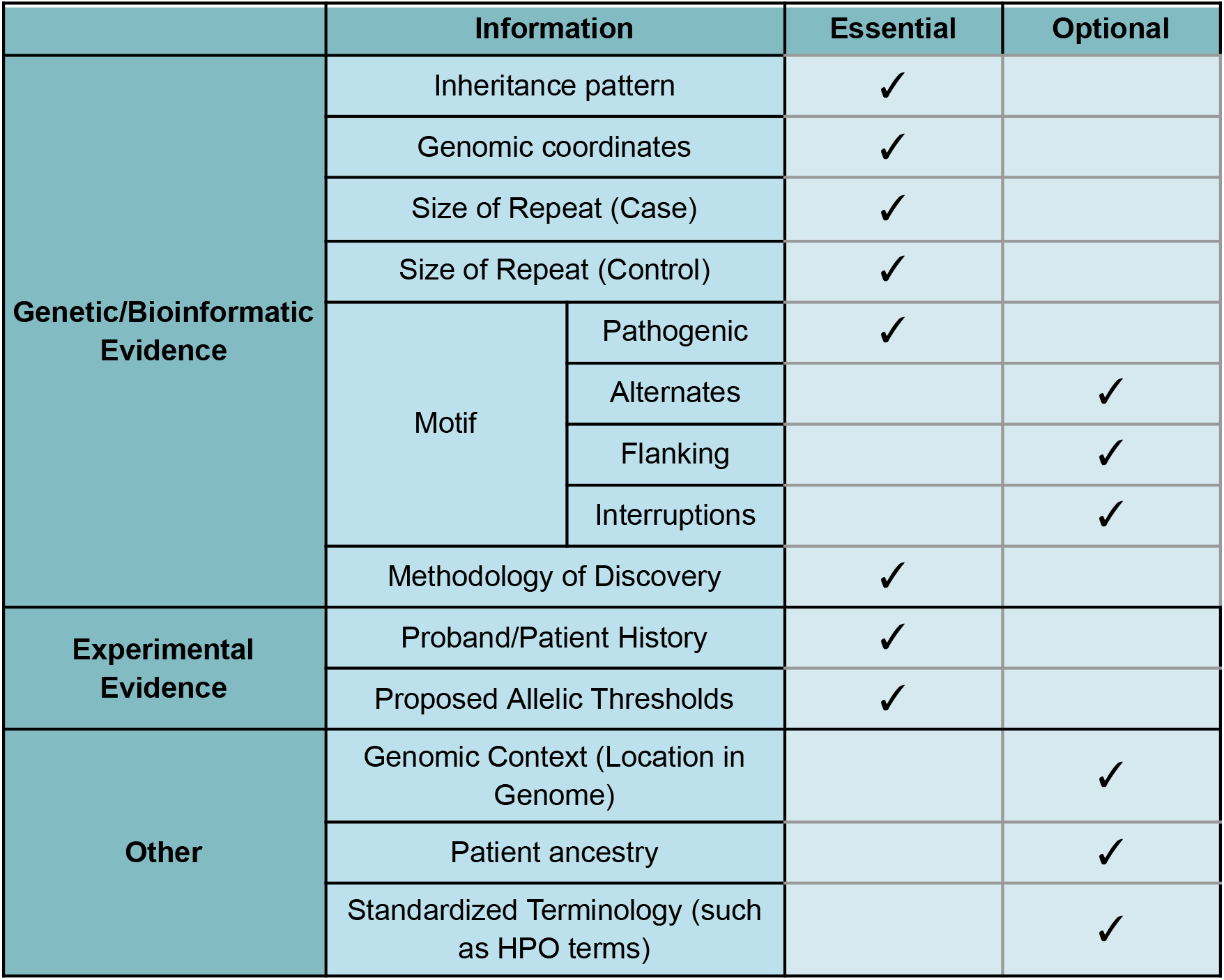
Proposed minimum evidence for reporting a new TR locus

Specifying an inheritance pattern is essential to this process as it allows scorers to evaluate segregation evidence and determine pathogenic details from coincidental findings. Genomic coordinates are often not cited directly, generally buried in figures, supplementary materials, or found through inferences from a mentioned intron or exon. Reporting the repeat length in both affected individuals and controls is important for establishing benign, intermediate, and pathogenic ranges. The exact sequence of the motif (including alternate motifs or interruptions) should be documented, as interruptions can influence stability or expression. The analytic approach should also be reported to allow assessment of accuracy and reliability, given the impact of methodological choices on TR detection and interpretation.

Our advised minimum clinical evidence includes proband/patient medical history and proposed allelic thresholds. Patients’ medical histories are important at many points in evaluating TRs, including determining clinical relevance of an analogous variant, correlations between allele size/composition and impact on disease phenotype (such as age of onset, severity, and clinical manifestations), and determining the associated phenotype. Reporting the frequency of symptoms within a cohort is also useful in determining and defining a phenotype. While not necessary, we also suggest some additional evidence for maximum clarity. For example, reporting the genomic context (e.g., exonic, intronic, non-coding) provides further localization information as well as mechanistic insight, expediting future research. Patient ancestry informs allele frequency interpretation and allows population-specific data to be aggregated. Finally, using standardized terminology (such as HPO terms) reduces ambiguity when describing patient phenotypes.

## Conclusions

The evaluation of tandem repeat locus-disease relationships has been limited by procedures designed for gene-level variants, which overlook the distinctive aspects and variability of TR loci. To address this gap we introduced criTRia: a standardized, repeat-aware scoring system that operates at the locus level rather than the gene level. By incorporating TR-specific evidence categories, reweighting scoring for experimental and genetic evidence, and updating rescoring intervals, criTRia captures the distinct complexities of repeat expansions and contractions. This framework also lays the groundwork for population-level frequency estimation of these rare diseases. Applying criTRia to 65 loci from STRchive^10^ demonstrated its capacity to accurately classify both well-established and emerging TR locus-disease relationships. Beyond improving consistency and transparency in curation, criTRia enhances diagnostic precision and supports the discovery of novel repeat-associated mechanisms. As TR research continues to expand, maintaining a TR-focused framework will be essential for broader clinical genetics and diagnostic practice. Future iterations of criTRia will benefit from integrating new genomic resources, population-level data, and mechanistic insights, ensuring that TR-specific scoring evolves alongside the field. Ultimately, criTRia establishes the foundation for a unified, evidence-based standard that will advance both clinical and research applications of tandem repeat genomics.

## Supporting information

Supplemental File 1. criTRia Standard Operating Procedure, Version 1

Supplemental File 2. Prompt for LLM-assisted evidence-detail generation

Supplemental File 3. Table of GenCC gene-level curation scores awarded to tandem repeat diseases, and criTRia locus-level curations

Supplemental File 4. Table of detailed criTRia TR disease-locus curations

## Data Availability

All scoring was performed using publicly available data, primarily those compiled in STRchive and from the scientific literature. Numerical and categorical scoring, as well as cross-group comparisons, were organized in Google Sheets and Microsoft Excel. All scripts for data processing, score aggregation, and figure generation were developed and written in Posit Cloud using R software. The scripts used to generate the main and supplemental files are publicly available at https://github.com/dashnowlab/criTRia. The criTRia Standard Operating Procedure and full scoring matrices are available in the Supplemental Files.

https://strchive.org/critria/

https://github.com/dashnowlab/criTRia

## Data Availability

All scoring was performed using publicly available data, primarily those compiled in STRchive and from the scientific literature. Numerical and categorical scoring, as well as cross-group comparisons, were organized in Google Sheets and Microsoft Excel. All scripts for data processing, score aggregation, and figure generation were developed and written in Posit Cloud using R software^36,37^. The scripts used to generate the main and supplemental files are publicly available at https://github.com/dashnowlab/criTRia. The criTRia Standard Operating Procedure and full scoring matrices are available in the Supplemental Files.

## Acknowledgments

The authors would like to thank Vincent Rubinetti for designing and developing the STRchive and criTRia website and providing feedback on data structure and automation. We thank Gabriel Zinser for providing curation assistance. We would also like to thank Alicia Byrne and Grace E. VanNoy for feedback during the initial conceptualization stage.

## Author Contributions

Contributions are described following the https://credit.niso.org/framework.

Macayla Ann Weiner: Conceptualization, Data Curation, Formal analysis, Software, Writing - Original Draft, Writing - Review & Editing, Visualization Laurel Hiatt: Conceptualization, Data Curation, Methodology, Writing - Original Draft, Writing - Review & Editing Pamela Ajuyah: Methodology, Writing - Review & Editing

Elbay Aliyev: Data Curation, Formal analysis, Software, Writing - Original Draft, Writing - Review & Editing Harriet Dashnow: Data Curation, Software, Writing - Original Draft, Writing - Review & Editing, Visualization, Supervision.

## Ethics Declaration

Not applicable.

## Declaration of generative AI and AI-assisted technologies in the writing process

During the preparation of this work, the author(s) used ChatGPT version GPT-5.5 Thinking (OpenAI) to assist with semi-automated generation of concise evidence-detail text for curated tandem repeat locus–disease entries. Grammerly was used to edit the manuscript for grammar. After using these tools, the authors reviewed and edited the content as needed and take full responsibility for the publication’s content.

## Supplemental Files

**Supplemental File 1**. criTRia Standard Operating Procedure, Version 1

**Supplemental File 2**. Prompt for LLM-assisted evidence-detail generation

**Supplemental File 3**. Table of GenCC gene-level curation scores awarded to tandem repeat diseases, and criTRia locus-level curations

**Supplemental File 4**. Table of detailed criTRia TR disease-locus curations

