## Supplemental File 1. criTRia Standard Operating Procedure, Version 1 for "criTRia: A Classification System and Evidence Criteria for Tandem Repeat Locus-Disease Relationships"

### criTRia Standard Operating Procedure, Version 1
### A Classification System and Evidence Criteria for Tandem Repeat Locus-Disease Relationships

Macayla Ann Weiner[1], Laurel Hiatt[2], Pamela Ajuyah[3], and Harriet Dashnow[1]

1. Department of Biomedical Informatics, University of Colorado Anschutz, Aurora, CO, USA
2. Department of Human Genetics, University of Utah, Salt Lake City, UT, USA
3. Program in Medical and Population Genetics, Broad Institute of MIT and Harvard, Cambridge, MA, USA

#### Table of Contents

**Background** 3
**Overview** 3
**Categorical Classifications** 4
**Evidence Collection** 7
Lumping and Splitting 7
**Minimum Evidence** 8
**Literature Search** 9
**Broad considerations for scoring** 9
**Scoring Evidence** 10
Scoring Matrix for all Evidence 10
Genetic/Bioinformatic Evidence 10
Genetic Evidence Scoring Matrix 12
Experimental Evidence 12
Experimental Evidence Scoring Matrix 15
**References** 16

#### Background

The criTRia tandem repeat locus curation process is a framework to evaluate the strength of a monogenic locus-disease relationship based on publicly available evidence. While established protocols exist for evaluating monogenic gene-disease relationships and variant interpretation[1–11], these do not account for the specific considerations of tandem repeat disease loci. In particular, we consider the evidence that a specific tandem repeat locus is associated with a disease rather than considering the gene as a whole. criTRia builds on ClinGen's established gene-disease relationship standard operating procedure[12], and adapts and extends it to specifically address tandem repeat diseases through tailored criteria and adjusted score weighting. Much of the procedure remains unchanged, ensuring consistency and only modifying what was necessary. It is important to note that there are emerging disease-associated loci within the same gene, some of which are very close to one another, and that these require special care.

#### Overview

The criTRia scoring process uses the following framework to assign a score and classification for a locus-disease relationship:

- Establishing the locus-disease mode of inheritance
- Evidence collection
    - Genetic Evidence
    - Experimental Evidence
- Evaluation and scoring of evidence
- Review, classification, and approval of a locus-disease relationship
- Publication of the final classification and supporting evidence
- Re-evaluation as indicated

In the subsequent sections of this document, each step will be outlined in detail, along with general recommendations.

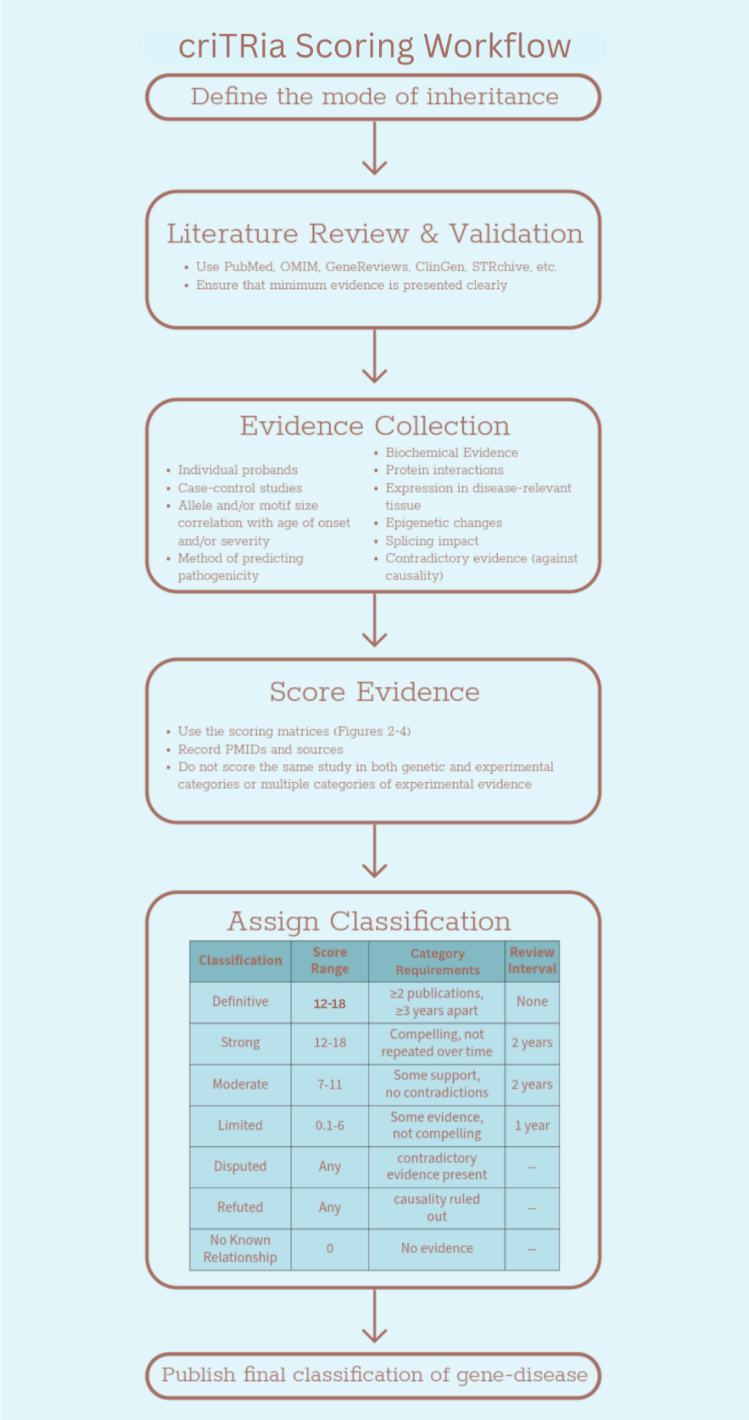


***Figure 1.*** *Flowchart detailing the steps for discovering and publishing new loci*.

#### Categorical Classifications

*Definitive*

- Score: 12-18
- Replicated over time (>3 years)

The role of the evaluated locus/gene in the evaluated disease has been repeatedly demonstrated in both research and clinical settings, and *<u>has been replicated over time</u>* (at least 2 publications with convincing evidence at least 3 years apart). Repeat expansions that disrupt gene function and are supported by strong genetic and population data (e.g, absence of the repeat expansion in study controls/unaffected individuals, *de novo* expansion, segregation with disease in families, or consistent linkage to a genomic region) are considered convincing of disease causality in this process.

- Interval for re-evaluation: no set time point
- Re-evaluation may be considered if contradictory information has arisen or a need has been expressed for another reason.

*Strong*

- Score: 12-18
- Not replicated over time (>3 years)

The role of the evaluated locus in the evaluated disease has been independently demonstrated in at least two separate studies, providing strong supporting evidence for this gene's role in the disease, although not validated over time. Locus-disease pairs with strong evidence demonstrate considerable genetic evidence (e.g., several unrelated probands harboring variants with sufficient supporting evidence for disease causality). Compelling evidence from different types of supporting experimental data is typically also present, but is not necessarily required to reach this designation if substantial convincing genetic evidence is present. In addition, no convincing evidence has emerged that contradicts the role of the locus in the noted disease.

- Interval for re-evaluation: two years after last scoring

*Moderate*

- Score 7-11

There is moderate evidence to support a causal role for the evaluated locus in the evaluated disease. Locus-disease pairs with moderate evidence typically demonstrate *some* convincing genetic evidence (e.g., unrelated probands harboring variants with sufficient supporting evidence for disease causality, with or without experimental data supporting the locus-disease relationship). The role of this locus in the disease may not have been directly reported, but no convincing evidence has emerged to contradict its role in the noted disease.

- Interval for re-evaluation: two years after last scoring

*Limited*

- Score: 0.1-6

In this category, there is limited evidence supporting the locus-disease relationship. The Limited category should not be applied in circumstances where none of the presented evidence is compelling; in these circumstances, the Disputed category should be considered.

- Interval for re-evaluation: one year after last scoring

*No Known TR Locus-Disease Relationship*

Evidence for a causal role in the monogenic disease of interest has not been reported within the literature (published, pre-published, and/or present in public databases, e.g., ClinVar). These genes might be “candidate” genes based on linkage intervals, animal models, and/or their involvement in pathways known to be involved in human disease, but no reports have directly implicated the gene or locus in the specified disease. If a claim of a relationship with the specified disease has been reported, but the evidence is minimal or not compelling, consider Limited, Disputed, or Refuted.

A notable difference in criTRia’s procedure compared to ClinGen’s: A tag designating “animal model only” is applied on [clinicalgenome.org](clinicalgenome.org) for those gene-disease pairs in which no human genetic evidence has been asserted, but an animal model exists.

*Contradictory*
A locus-disease pair may be classified as Disputed or Refuted when significant contradictory evidence emerges, or the strength of previously reported supporting evidence diminishes. While genetic and experimental data may initially support the relationship, conflicting evidence calls the association into question. The choice between these classifications is based on the strength of the data. It is best to use Disputed when supporting and contradictory evidence are both compelling. Refuted should be used when the contradictory evidence substantially outweighs any supporting evidence. Expert input is essential when dealing with contradictory evidence and when using one of these classifications. More details about these categories and example scenarios are described below.

<u>Disputed</u>:

- The presented clinical cases have disparate phenotypes.
    - Notably distinct phenotypes between cases, such as phenotypes in completely different patient systems (neuromuscular versus respiratory symptoms)
    - Negligible overlap in symptoms between cases, such that cases are not convincingly connected
- Proposed pathogenic variants are observed in unaffected individuals or control populations at a rate higher than expected based on the disease prevalence and proposed penetrance.
- There are cases in the research with unclear or differing modes of inheritance.
- <u>*Alternative causes have NOT been identified in the original proband, while there is contradictory evidence.*</u>

<u>Refuted</u>:

- The evidence against the locus-disease relationship outweighs the evidence for it, or the locus-disease relationship and its previous evidence have been ruled out.
- <u>*Alternative causes for the disease are identified in the original proband.*</u>
- The proband is later determined not to have the disease in question.
- Statistically rigorous case-control shows no enrichment.

**<u>Evidence Collection</u>**

Evidence is collected primarily from published peer-reviewed literature, but can also be present in publicly accessible resources, such as variant databases, and high-quality pre-published literature (preprints) with compelling evidence used with discretion in scoring. When determining whether a case is appropriate for use, consider the following:

- Sufficient evidence is provided.
- The case must be publicly accessible. For example, do not include cases that are only available to authorized users.
- The case is well-described with appropriate phenotype, methodology, statistical analysis when appropriate, and other variant information.
- Case is not otherwise believed to be described in already scored literature – for example, if a proband is included across multiple studies, only score them once.
- If a locus is proposed as pathogenic for the first time, the case must have supporting evidence that the variant is causative beyond just its presence in an affected individual.
- Previous gene curation is not a prerequisite. As we have seen, some TRs are clearly pathogenic, but there are no other variant types in the gene associated with the disease.
    - I.e., Huntington's Disease (*HTT*) or Myotonic Dystrophy 1 (*DMPK*)

Evidence that has an associated identifier is preferred, typically a PMID. Check whether the database includes citations with PMIDs, as this will allow you to submit evidence more easily, and submit the DOI of anything without.

**Lumping and Splitting**

Give special care to cases where multiple tandem repeat loci are reported in the same gene. Many such loci have only recently been identified, and, in turn, much of the literature on these loci may not be as locus-specific as presented. Unfortunately, these issues vary greatly, so there is no singular approach to resolve all complexities. Instead, we urge scorers to note phenotypic and bioinformatic evidence and to use their best judgment. Isolating evidence for specific loci within a gene's phenotypic spectrum may be determined by symptoms or age of onset. Evidence from TR loci on the same gene should be defined by coordinate; however, when that information is unavailable or impractical, distinct loci may be defined by the motif, pathogenic repeat range, or other positional clues. A potential option is to score experimental evidence as it applies, and to exclude non-specific genetic data. The following examples have been included to aid in deciding how to approach such diseases.

<u>Hand-foot-genital (HFG) syndrome of *HOXA13*</u>: This phenotype has been linked to tandem repeat expansions in three distinct polyalanine tracks, each separated by ~60 base pairs. This has led to the proposed three conditions: HFG-I, HFG-II, and HFG-III associated with the loci *HOXA13-I, HOXA13-II,* and *HOXA13-III*, respectively. For most papers, the pathogenic range of the repeat provided a clear indication of which type of HFG the publications studied. Any literature that remained unclear was scored across all three loci, restricted to the experimental evidence category as applicable to all three HFG types. It should be noted that the pathogenic repeat range is usually not a viable option and should be used as a last resort in separating intragenic disease loci.

Diseases of *FMR1* (gene): Fragile X syndrome (FXS), fragile X-associated tremor/ataxia syndrome (FXTAS), and fragile X-associated primary ovarian failure (POF1) present a phenotypic spectrum within the same gene, *FMR1*. Identifying FXS-focused papers was fairly straightforward, as most defined the FXS-specific repeat range. It became increasingly clear that FXTAS and POF1 were intertwined in the literature, as there were overlapping symptoms in XX individuals with both diseases and similar bioinformatic evidence. For this reason, we decided to group FXTAS and POF1 and score FXS separately.

The *ARX* phenotypic spectrum: Expansions in *ARX* vary in length and location on the gene and cause a range of diseases. Partington syndrome (PRTS) and early infantile epileptic encephalopathy (EIEE1) were the focus of our curation and scoring, given previous associations with TRs. Most studies were easily assigned to one disease, but for those that were not, experimental evidence was scored and attributed to both diseases if the findings were relevant to both phenotypes. It is worth noting that attempts to define diseases based on coordinates were not particularly fruitful, as even the same locus can exhibit variable phenotypes and disease associations.

**Minimum Evidence**

We strongly urge scorers and researchers to ensure that sufficient evidence is provided for a newly presented locus-disease relationship. Documenting and reporting this evidence will allow for future replication and clarity. Following is the minimum genetic and bioinformatic evidence we propose for inclusion:

- Proposed inheritance pattern (e.g., autosomal recessive or X-linked dominant)
- Genomic coordinates
  - Chromosome, start, end, and reference genome build
  - For both controls and cases, coordinates should be explicit or straightforward to derive
- Size in repeats of all alleles analyzed (control and case)
- Motif of repeat, including alternate motifs/interruptions and flanking repeats (if any)
  - Are there distinct motifs present in the data?
  - What is the role of interruptions?
- Methodology of discovery/validation
  - Sequencing technology
  - Genotyping tool
  - Any molecular approach to orthogonally validate alleles

Our minimum clinical evidence includes:

- Proband/patient medical history
  - Age of onset for first symptoms
  - Familial history of disease (positive or negative)
  - Frequency of symptoms, if the study includes a cohort
- Proposed allelic thresholds (benign and pathogenic ranges)

- Benign ranges may be based upon population data or segregation analysis, where there are sufficient controls, while pathogenic thresholds are often defined as the size at which disease manifests
  - Molecular testing can be used to observe functional changes to refine pathogenic thresholds further

We also suggest the following evidence (although it is not as crucial as the above:

- Genomic context (coding, 5'UTR, etc.)
- Gene strand
- Proband/patient ancestry
- HPO terms

#### Literature Search

The "Literature Search" section in ClinGen's most recent SOP[12] provides extensive guidance on conducting a targeted, effective literature search for gene-disease relationships, which is useful for broader searches. We recommend that the scorer use their best professional judgment to ensure they also find research on locus-phenotype relationships, specific to the TR context. After a broad search, it may be useful to narrow it with additional terms such as "tandem repeat" or "repeat expansion". For a list of useful websites, see Appendix A in ClinGen's most recent SOP[12].

#### Broad considerations for scoring

The default scoring of a piece of evidence can be increased by the strength of the evidence, as well as the addition of multiple pieces of evidence within the same publication. For instance, the default score of 0.5 for a proven biochemical alteration may be increased to 1 if additional protein characterization is included in the study. Scorers should aim to score a range of publications, where possible, and ensure they are independent. Ensure that scoring considers the section's maximum score as well as the maximum overall score for the evidence category. Evidence should not be double-counted. A publication may include multiple pieces of evidence, but each empirical observation (e.g., a specific experiment or observation) may contribute to only a single evidence category. This prevents inflation of evidence strength and improves reproducibility across curators. Genetic evidence should receive priority over experimental evidence. For example, if a study with a featured proband includes a patient cell line and a non-human model, the cell line evidence may be used to increase the genetic evidence score while the model organism is cataloged as experimental evidence. The patient cells category of experimental evidence would thus require a separate study/proband characterization for evidence to be scored there. However, if sufficient information is available, allele size/anticipation data, case-control information, and segregation evidence can be obtained from the same study in which probands are evaluated.

The highest priority in cataloging evidence should be given to locus-specific studies that establish an association between the TR variant and the clinical phenotype. Still, gene-level experimental findings can be informative in demonstrating the functional consequences of alterations in the region. Evidence that can be attributed to a gene but not to a specific tandem repeat locus should be scored as experimental evidence and not as genetic evidence.

Evidence that the reviewer finds to be specious or questionable should be scored as 0 when documented, regardless of evidence type. Such evidence may lead to a limited or disputed classification.

**Scoring Evidence**

The scores of the genetic and experimental evidence categories are summed. Total evidence may sum to a maximum of 18 (12 points of genetic evidence and 6 of experimental evidence), with maximum scores within each evidence section as well. We suggest prioritizing genetic evidence over experiments in collection, as it is often specific to TR disease considerations and can be sufficient for a Definitive classification on its own. It is important to document the evidence sources (PMIDs, ClinVar IDs, etc.). If evidence came from a review article or an overview resource such as OMIM or GeneReviews, cite both the review and the original research. While we do offer default scores, we suggest an increase if there are multiple forms of evidence for the same category. Examples include having multiple animal cell models or evidence of association between allele length/motif and age of onset, penetrance, and/or severity. A piece of evidence that is especially convincing is not grounds for increasing the score, as there is no way to standardize this. It is also important to make note of why a certain score was given (i.e., “Study details two animal models, so a score of 4 was granted”).

#### Scoring Matrix for all Evidence

| | Evidence Category | Evidence Type | Description | Default Score | Suggested Upgrades | Scoring Range | Maximum Score | Maximum Category Score |
|---|---|---|---|---|---|---|---|---|
| Genetic Evidence | Singular Evidence | Probands | Unrelated Probands | 0.5 | +0.5 for inheritance/de novo evidence, +0.5 for functional evidence | 0-1.5 points per proband | 6.0 | 6.0 |
| | Collective Evidence | Allele | Relationship between allele and/or motif vs. age of onset and/or severity | 1.0 | Multiple forms of evidence: I.e Motif length affects both age of onset and severity | 0-2 | 2.0 | 3.0 |
| | | Computational | Computational Pathogenicity Prediction | 0.5 | Multiple methods of prediction | 0-1.5 | 3.0 | |
| | | Segregation | Linkage Region for [illegible] | 1.5 | -- | -- | 1.5 | |
| | Statistics | Case-Control Data | 1. Variant Detection Methadology<br>2. Power<br>3. Bias and Confounding<br>4. Statistical significance | -- | 6: appropriately matched case and controls, no biases/confounding factors, highly statistically significant<br>4: no appropriately matched, p-value mdoeratety statistically significant<br>2: aggregate/population data, p-value not very significant<br>0: detection method differs (may be case-level) | 0-6 | 12.0 | 12.0 |
| | Maximum Overall Score for all Genetic Evidence | | | | | | | 12.0 |
| Experimental Evidence | **Evidence Category** | **Evidence Type** | **Description** | **Default Score** | **Suggested Upgrades** | **Scoring Range** | **Maximum Score** | **Maximum Category Score** |
| | Function | Biochemical function | Enzymatic activity, DNA/RNA binding, structural roles, etc. | 0.5 | Multiple foms of evidence | 0-2 | 2.0 | 2.0 |
| | | Protein Interaction | Protein product interacts with other disease-relevant proteins or complexes. | 0.5 | Multiple foms of evidence | 0-2 | 2.0 | |
| | | Regulatory Impact | Including gene expression, epigenetic changes, or impacts on splicing. | 0.5 | Multiple foms of evidence | 0-2 | 2.0 | |
| | Functional Alteration | Patient Cells | Patient cells display disease-relevant phenotypes | 1.0 | Multiple patient cells studied in different experiments | 0-2 | 2.0 | 2.0 |
| | | Non-patient Cells | Test the impact of candidate repeat variants in controlled conditions. | 0.5 | Multiple control sample cells studied in different experiments | 0-1 | 1.0 | |
| | Models | Non-Human Model Organism | Used to support genetic and phenotypic findings and should mimic the environment of the diseased human tissue | 2.0 | Multiple models studied: I.e. Mouse and zebrafish models in the same experiment | 0-4 | 4.0 | 4.0 |
| | | Cell Culture | | 1.0 | Multiple different types of cells cultured | 0-2 | 2.0 | |
| | Rescue | Human treatment | Restoring gene function reverses disease phenotypes in humans | 2.0 | Rescues in multiple different healthy human cells | 0-4 | 4.0 | |
| | | Non-Human Model Organism | [illegible] phenotypes by removing the repeat, restoring gene expression, expressing a normal allele or silencing the [illegible] | 2.0 | Rescues in multiple moodels: I.e. Mouse and zebrafish | 0-4 | 4.0 | |
| | | Cell Culture | | 1.0 | Rescues in different types of cultured cells | 0-2 | 2.0 | |
| | | Patient Cells | | 1.0 | Rescues in multiple difernt patient cells or methods of rescue | 0-2 | 2.0 | |
| | | Maximum Overall Score for All Experimental Evidence | | | | | | 6.0 |

***Figure 2.*** *Scoring matrix for all evidence, genetic and experimental.*

##### Experimental Evidence

There are several assay methods used to elucidate gene function. For clinical validity classifications, only evidence that supports the role of a locus in a disease, or phenotypic features related to the disease entity of interest, should be scored. Expert panels should identify validated functional assays or, if they are curator-identified, confirm them by expert review.

*Functional Evidence*

Biochemical Function

This segment catalogs evidence that the gene or repeat locus has a known molecular or cellular function relevant to the disease process. This may include enzymatic activity, DNA/RNA binding, transcriptional regulation, or structural roles. Disruption of this function by repeat expansion or mutation is likely to support pathogenicity. This segment has a range of 0–2 with a default score of 0.5. Suggested upgrades may be multiple forms of evidence (i.e., both RNA binding and transcriptional regulation).

Protein Interaction
Data show that the protein product of the gene interacts with other disease-relevant proteins or complexes. Repeat expansions that affect these interactions—either by disrupting binding or creating toxic interactions—support a functional role in disease. This segment ranges from 0 to 2, with a default score of 0.5, which can be increased if multiple experimental methods are used to assess protein interactions or if compelling simulations leverage computational strategies.

Regulatory Impact
Evidence that the repeat expansion or contraction has a substantial regulatory impact, including data reflecting altered gene expression, epigenetic changes, or abnormal splicing. Expression evidence may also demonstrate that the gene is expressed in disease-relevant tissues, or that expression levels change with the size of the repeat expansion. Evidence of epigenetic changes may indicate that the repeat expansion triggers epigenetic modifications, such as DNA methylation, histone modifications, or chromatin remodeling. These may result in transcriptional silencing or altered expression patterns. Splicing impact may be shown by evidence demonstrating that a repeat expansion alters pre-mRNA splicing, leading to exon skipping, intron retention, or cryptic splice-site use. RT-PCR, RNA-seq, or splice reporter assays may show this. This category ranges from 0-2, with a default score of 0.5 and a suggested upgrade for multiple forms of evidence supporting it.

*Evidence of Functional Alteration*
Patient Cells
Functional studies using cells derived from affected individuals. These reflect disease-relevant genotypes and can show expression changes, aggregation, or other molecular phenotypes. This segment ranges from 0 to 2, with a default score of 1, which can be upgraded if there are multiple patient cell types (e.g., hiPSCs and living patient cells).

Non-Patient Cells
Studies in cell lines that do not originate from patients, but are used to test the impact of repeat expansions or candidate variants in controlled conditions. This segment ranges from 0 to 2, with a default score of 0.5. This may be upgraded when multiple cell lines are used in the study, while still explicitly excluding cell culture (see Cell Culture Model below). A prominent example is when there are multiple control groups, such as using unaffected family members and unaffected subject data from a database.

*Model-based Evidence*
Evidence in this category should support genetic and phenotypic findings and mimic the environment of diseased human tissue[13].

Non-human Model Organism
*In vivo* models are used to test the effects of repeat expansions or gene loss/gain. These models may show behavioral, anatomical, or molecular phenotypes that correspond to human disease. This segment ranges from 0 to 4, with a default score of 2 per model (e.g., fly and mouse).

<u>Cell Culture Model</u>
Any *in vitro* cell-based assay, including patient-derived, immortalized, or stem-cell-derived models. Cell Culture Models differ from the Cell categories above in that they recapitulate features of the disease tissue environment. Generally used to study molecular mechanisms, expression, and toxicity in a controlled setting. This segment ranges from 0 to 2, with a default score of 1.

*Rescue Evidence*
Reflects instances in which phenotypic rescue is performed, either genetically or molecularly. Suggested upgrades for the following evidence categories include rescues across multiple cell types or multiple rescue strategies for the same cell type.

<u>Human Treatment</u>
Evidence that restoring gene function reverses disease phenotypes in human controls. This is strong evidence of causality but rarely available; therefore, this segment has a range of 0-4 with a default score of 2.

<u>Non-human model organism</u>
Reversal of disease-like phenotypes in a non-human model by removing the pathogenic allele, restoring gene expression, or expressing a normal allele. Supports the functional relevance of the locus and repeat. This segment ranges from 0 to 4, with a default score of 2.

<u>Cell Culture Model</u>
Evidence that introducing a normal gene or silencing the mutant allele in a cell line can reverse pathological features like aggregation, toxicity, or mis-splicing. This segment ranges from 0 to 2, with a default score of 1.

<u>Patient Cells</u>
Similar to above, but specifically in cells derived from affected individuals. Rescue may involve gene editing, knockdown of repeat-containing transcripts, or antisense approaches to restore cellular function. This segment ranges from 0 to 2, with a default score of 1.

| | Evidence Category | Evidence Type | Description | Default Score | Suggested Upgrades | Scoring Range | Maximum Score | Maximum Category Score |
|---|---|---|---|---|---|---|---|---|
| Experimental Evidence | Function | Biochemical function | Enzymatic activity, DNA/RNA binding, structural roles, etc. | 0.5 | Multiple foms of evidence | 0-2 | 2.0 | 2.0 |
| | | Protein Interaction | Protein product interacts with other disease-relevant proteins or complexes. | 0.5 | Multiple foms of evidence | 0-2 | 2.0 | |
| | | Regulatory Impact | Including gene expression, epigenetic changes, or impacts on splicing. | 0.5 | Multiple foms of evidence | 0-2 | 2.0 | |
| | Functional Alteration | Patient Cells | Patient cells display disease-relevant phenotypes | 1.0 | Multiple patient cells studied in different experiments | 0-2 | 2.0 | 2.0 |
| | | Non-patient Cells | Test the impact of candidate repeat variants in controlled conditions. | 0.5 | Multiple control sample cells studied in different experiments | 0-1 | 1.0 | |
| | Models | Non-Human Model Organism | Used to support genetic and phenotypic findings and should mimic the environment of the diseased human tissue. | 2.0 | Multiple models studied: I.e. Mouse and zebrafish models in the same experiment | 0-4 | 4.0 | 4.0 |
| | | Cell Culture | | 1.0 | Multiple different types of cells cultured | 0-2 | 2.0 | |
| | Rescue | Human treatment | Restoring gene function reverses disease phenotypes in humans | 2.0 | Rescues in multiple different healthy human cells | 0-4 | 4.0 | |
| | | Non-Human Model Organism | Reversal of disease-like phenotypes by removing the repeat, restoring gene expression, expressing a normal allele or silencing the mutant allele. | 2.0 | Rescues in multiple moodels: I.e. Mouse and zebrafish | 0-4 | 4.0 | |
| | | Cell Culture | | 1.0 | Rescues in different types of cultured cells | 0-2 | 2.0 | |
| | | Patient Cells | | 1.0 | Rescues in multiple difernt patient cells or methods of rescue | 0-2 | 2.0 | |
| | | | | | Maximum Overall Score for All Experimental Evidence | | | 6.0 |

**Experimental Evidence Scoring Matrix**

***Figure 4.*** *Scoring matrix for Experimental Evidence*

**Genetic Evidence**

Genetic evidence may be derived from singular data (studies describing individuals or families with variants in the gene of interest), collective evidence, and/or statistical data (studies in which statistical analysis is used to evaluate enrichment of variants in cases compared to controls), but case-control data is preferred. While a single publication may include all three, individual cases should not exceed the section's maximum scores. For example, if a case from a case-control study were singled out for detailed discussion in the publication and familial inheritance and pedigree information were provided, this case could be assigned the maximum score and would not be used for any further evidence. The scorer should determine which is the stronger piece of evidence and include those in the curation.

*Singular Evidence*

Proband evidence from case or family studies: Scoring is per unrelated proband, so a family with multiple affected individuals can only have one proband selected for scoring. The data may provide evidence of inheritance or function, but this is not required to receive a score. This segment has a range of 0-6 total with 0.5 awarded for each proband; an additional 0.5 if there is functional data; and an additional 0.5 if parent alleles were assessed and are consistent with the mode of inheritance—whether the variant is *de novo*, or other such familial information—for a total of 1.5 possible for each proband/family. The score may be upgraded per unrelated proband.

*Collective Evidence*

Repeat Length/Motif Consequences:

Data show relationships between allele size and age of onset, anticipation, and/or between allele size and disease severity. This segment has a range of 0–2 with a default of 1. A potential scenario for an increase in score is a study presenting evidence of correlations between age of onset and disease severity and allele size.

Computational Pathogenicity:
Predicted pathogenicity reflects evidence that leverages computational predictive strategies to infer the likelihood of pathogenicity for the locus. Such evidence may be assessed using appropriate constraint metrics or tandem-repeat specific tools such as RExPRT or SISTR[814]. Recent studies have proposed that the degree of repeat polymorphism in the general population is correlated with pathogenicity[15]. This segment ranges from 0 to 1.5, with a default score of 0.5. Suggested upgrades include evidence of multiple of the stated assessment measures[16].

Segregation
Evidence can be supported by a linkage region overlapping the candidate locus and coinciding with disease status within families, as indicated by LOD scores. A score of 1.5 is given to any study with a significant LOD score. For information about calculating an LOD score, please see the Segregation Analysis section of ClinGen's most recent SOP[12].

*Statistical Data*
Case-Control Data
This segment ranges from 0 to 6. Data that receives the maximum score should have appropriately matched cases and controls, with no biases or confounding factors, and should be highly statistically significant. Experiments where the samples are not appropriately matched, and the p-value is moderately significant, should receive a score of 4. Data using aggregate or population databases with a hardly significant p-value should be awarded 2. In any experiment in which the method of detection differs (such as case-level data), 0 should be awarded. Four types of evidence should be examined:

1. Variant Detection Methodology: Short-read or long-read sequencing, targeted PCR-based assays, or bioinformatic tools that infer repeat sizes from alignment patterns.
2. Power: The ability of a study to detect a true association between a genetic variant and a phenotype, depending on sample size, effect size, allele frequency, and phenotype specificity.
3. Bias and Confounding: Systematic errors that skew the results, or the presence of external variables that distort the true relationship between the variant and disease.
4. Statistical Significance: A significant p-value is reported in the literature.

**Genetic Evidence Scoring Matrix**

| | Evidence Category | Evidence Type | Description | Default Score | Suggested Upgrades | Scoring Range | Maximum Score | Maximum Category Score |
|---|---|---|---|---|---|---|---|---|
| Genetic Evidence | Singular Evidence | Probands | Unrelated Probands | 0.5 | +0.5 for inheritance/de novo evidence, +0.5 for functional evidence | 0-1.5 points per proband | 6.0 | 6.0 |
| | Collective Evidence | Allele | Relationship between allele and/or motif vs. age of onset and/or severity | 1.0 | Multiple forms of evidence: I.e Motif length affects both age of onset and severity | 0-2 | 2.0 | 3.0 |
| | | Computational | Computational Pathogenicity Prediction | 0.5 | Multiple methods of prediction | 0-1.5 | 3.0 | |
| | | Segregation | Linkage Region for disease | 1.5 | -- | -- | 1.5 | |
| | Statistics | Case-Control Data | 1. Variant detection methodology<br>2. Power<br>3. Bias and Confounding<br>4. Statistical significance | -- | 6: appropriately matched case and controls, no biases/confounding factors, highly statistically significant<br>4: no appropriately matched, p-value mdoerately statistically significant<br>2: aggregate/population data, p-value not very significant<br>0: detection method differs (may be case-level) | 0-6 | 12.0 | 12.0 |
| | | | | | | | **Maximum Overall Score for all Genetic Evidence** | **12.0** |

***Figure 3.*** *Scoring matrix for Genetic/Bioinformatic Evidence* (link to full-size figure)


**Supplementary**

https://clinicalgenome.org/site/assets/files/10876/gene_disease_validity_standard_operating_procedures-_version_12.pdf

**S1.** ClinGen SOP 12

https://docs.google.com/spreadsheets/d/1VEuZqvwtQWzVSBc7Aj4FUPQABwyNTCcb8XDNCZ-xoKM/edit?usp=sharing

**S2.** All matrices included in this SOP and the associated manuscript
