## Supplemental File 2. Prompt for LLM-assisted evidence-detail generation for "criTRia: A Classification System and Evidence Criteria for Tandem Repeat Locus-Disease Relationships"

Macayla Ann Weiner[1], Laurel Hiatt[2], Pamela Ajuyah[3], Ebay Aliyev[1], and Harriet Dashnow[1]

1. Department of Biomedical Informatics, University of Colorado Anschutz Medical Campus, Aurora, CO, USA
2. Department of Human Genetics, University of Utah, Salt Lake City, UT, USA
3. Program in Medical and Population Genetics, Broad Institute of MIT and Harvard, Cambridge, MA, USA

**Supplementary Note**

**Standardized prompt for AI-assisted evidence-detail generation**

To support consistency and reproducibility across criTRia curations, we used a standardized prompt for AI-assisted generation of evidence-detail text. The same prompt structure was applied across curated tandem repeat locus–disease entries. ChatGPT was used only to draft concise evidence-detail descriptions from curator-provided materials, including source publications, partial criTRia JSON entries, and optional citation/source mappings. The prompt explicitly instructed the model not to modify evidence scores, add or delete evidence rows, alter evidence categories, change summary scores or classifications, or modify the JSON schema. All AI-assisted evidence-detail text was subsequently reviewed against the source literature and approved by at least one additional human reviewer before inclusion.

**AI-assisted evidence-detail generation prompt:**

"*I am uploading:*
*1. One or more paper PDFs*
*2. A partial criTRia JSON entry*
*3. Optional: PMID-to-paper/source mapping or curator notes*

*Task:*
*Act as a clinical geneticist/curator using criTRia SOP and scoring matrix. Read the uploaded paper(s) and update ONLY:*
*1. Existing "Evidence detail" fields that are null, vague, incomplete, or too informal.*
*2. The root-level "Description" field.*

*Important rules:*
*- Do NOT change any scores.*
*- Do NOT add new evidence rows.*
*- Do NOT delete evidence rows.*
*- Do NOT change category_summary, supercategory_summary, total_score, classification, publication_count, or publication_interval_years.*
*- Preserve the original JSON schema and field names exactly.*
*- Keep "Evidence detail" concise, factual, and directly tied to the assigned Evidence type and Score.*
*- If evidence is gene-level but not tandem-repeat/locus-specific, state this clearly in the Evidence detail.*
*- Do not double count the same proband, family, dataset, experiment, or observation.*
*- If an existing evidence row cannot be supported by the uploaded paper(s) or provided source mapping, keep the row but write: "Curator review needed: [brief reason]."*

*Handling multiple PMIDs / papers:*

*- Each evidence row has its own "Citation" field. Use the cited PMID/source in that row to match it to the correct uploaded PDF or provided source mapping.*
*- If multiple PDFs are uploaded, first identify which PDF corresponds to each PMID or citation.*
*- If I provide a PMID-to-paper/source mapping, use it to decide which source supports each evidence row.*
*- If an evidence row cites a PMID that is not represented by any uploaded paper or mapping, do not infer evidence from the PMID alone. Keep the row and write: "Curator review needed: cited source not provided."*
*- If one evidence row cites multiple PMIDs, use only the uploaded/provided sources that are available and clearly state which PMID/source supports the updated detail.*
*- If the uploaded paper supports only gene-level evidence and not tandem-repeat/locus-specific evidence, state this clearly in the Evidence detail and Evidence source report.*

*Return in the chat:*
*A. Updated JSON block*
*B. Evidence source report*
*C. Short curator-review note*

*Also create output files:*
*1. Updated JSON file:*
*- File name format: <Gene>_<Disease_ID>_updated.json*
*2. Excel report file:*
*- File name format: <Gene>_<Disease_ID>_evidence_source_report.xlsx*

*Do not provide long explanations unless there is uncertainty.”*
